# Continuous non-invasive hemodynamic monitoring in early onset severe preeclampsia

**DOI:** 10.1101/2023.04.10.23288375

**Authors:** Christina M Ackerman-Banks, Jasjit Bhinder, Maxwell Eder, Paul Heerdt, Lissa Sugeng, Jeffrey Testani, Aymen Alian, Heather Lipkind, Eric Velazquez, Uma Reddy, Josephine C Chou

## Abstract

**Objectives:** Continuous hemodynamic monitoring offers the opportunity to individualize management in severe preeclampsia (PEC). We compared cardiac output (CO) and systemic vascular resistance (SVR) measured by bioreactance (NICOM), Clearsite™ Fingercuff [CS), and 3D-echocardiography (3DE).

**Study Design:** This prospective observational study included 12 pregnant patients with early PEC. CO and SVR were measured simultaneously by NICOM, CS, and 3DE antepartum and 1-2 days postpartum. Using 3DE as the standard, CS and NICOM interchangeability, precision, accuracy, and correlation were assessed.

**Results:** Compared to 3DE-CO, CS-CO was highly correlated (R^2^=0.70, p=<0.0001) with low percentage error (PE 29%) which met criteria for interchangeablity. CS-SVR had strong correlation (R^2^=0.81, p=<0.0001) and low PE (29%). While CS tended to slightly overestimate CO (bias +2.05 +/-1.18 L/min, limit of agreement (LOA) -0.20-4.31) and underestimate SVR (bias -279 +/-156 dyes/sec/cm^5^; LOA -580-18.4) these differences were unlikely to be clinically significant. Thus CS could be interchangeable with 3DE for CO and SVR. NICOM-CO had only moderate correlation with 3DE-CO (R^2^=0.29, p=0.01) with high PE (52%) above threshold for interchangeability. NICOM-CO had low mean bias (−1.2 +/-1.68 L/min) but wide 95% LOA (−4.41 - 2.14) suggesting adequate accuracy but low precision in relation to 3DE-CO. NICOM-SVR had poor correlation with 3DE-SVR (R^2^=0.32, p=0.001) with high PE (67%), relatively low mean bias (238 +/-256), and wide 95% LOA (– 655–1131). NICOM did not meet the criteria for interchangeable with 3DE for CO and SVR.

**Conclusions:** Clearsite Fingercuff, but not NICOM, has potential to be clinically useful for CO and SVR monitoring in severe preeclampsia.

## Introduction

Hypertensive disorders of pregnancy (HDP) affect 2-8% of all pregnancies and have been increasing in frequency by up to 25% over the past two decades.^1,2^ It is a major contributor to maternal and neonatal severe morbidity and mortality with up to 16% of maternal deaths due to HDP.^3^ Early onset preeclampsia with severe features (PEC) is new hypertension with evidence of end-organ damage prior to 34 weeks gestation.^4^ It accounts for 20% of all cases of preeclampsia and is associated with more severe maternal and neonatal disease.^5^ Early onset PEC also has a distinct hemodynamic profile which manifests as lower cardiac output (CO) and higher systemic vascular resistance (SVR) as compared to both mild PEC and normal pregnancy.^6^ These abnormal hemodynamics may be predictive of maternal and fetal complications, such as severe hypertension, fetal growth restriction and fetal death.^5^ Continuous hemodynamic monitoring in severe PEC offers the potential to tailor antihypertensive therapy based on hemodynamic profiles, identify disease earlier, and ultimately improve maternal and fetal outomes.^6,7^ However, hemodynamic assessment in pregnancy has traditionally been difficult. Measuring invasive hemodynamics by Swan-Ganz catheter in pregnancy is neither feasible nor safe. Thus, non-invasive hemodynamics have usually been obtained with echocardiography. Unfortunately, this modality is time-consuming, operator dependent, and only available in discrete time intervals. Newer methods of continuous non-invasive hemodynamics through bioreactance (NICOM, Baxter Medical) or finger cuff pressure (Clearsite™[CS], Edwards) have been validated against invasive and non-invasive hemodynamics in other clinical settings, including normal pregnancy.^8-10^ But neither have been validated in pregnant patients with cardiac disease, including severe PEC. We sought to validate CO derived from NICOM (NICOM-CS) and Clearsite (CO-CS) against 3D echocardiography (3DE) derived CO (3DE-CO) in early onset severe PEC.

## Methods

### Study population

The Yale School of Medicine Human Investigation Committee approved this study. All patients signed an informed consent prior to enrollment. We prospectively enrolled 12 pregnant patients admitted with early onset PEC with severe features at a single center from December 2020 to October 2021. PEC with severe features was diagnosed clinically in accordance with diagnostic criteria by the American College of Obstetricians and Gynecologists.^4^ Inclusion criteria were singleton gestations with PEC with severe features diagnosed between 20 and 34 weeks gestation. Exclusion criteria included chronic hypertension requiring medications, age less than 18 years, and non-English primary language. Chronic hypertension was defined as systolic blood pressure(SBP) ≥ 140 mmHg and/or a diastolic blood pressure (DBP) ≥ 90 mmHg prior to 20 weeks gestation, as defined by the American College of Obstetricians and Gynecologists.^11^

### ClearSite FingerCuff (Edwards)

The CS Finger Cuff uses the Edwards EV1000 clinical platform which monitors hemodynamic parameters derived from continuous noninvasive measurement of the arterial pressure waveform. The finger cuff has a built in plethysmograph sensor to noninvasively measure the continuous finger arterial blood pressure using an inflatable cuff around the middle phalange of the finger. The brachial arterial pressure waveform is then reconstructed from the measured finger blood pressure pulsations to monitor systolic, diastolic, and mean arterial pressures (MAP). Hemodynamic parameters including CO, stroke volume (SV), and heart rate are calculated using a pulse contour method via the ClearSite Algorithm. SVR is calculated after the user manually entered a central venous pressure (CVP) value. These hemodynamic values are reported as 20 second averages on the monitor. The Heart Reference Sensor is used to compensate for hydrostatic pressure differences due to changes in height of the finger relative to the heart, with one end placed at the level of the patient’s finger and the other at heart level.^12^

### Noninvasive Cardiac Output Monitor (NICOM, Cheetah)

The Cheetah NICOM system is a noninvasive cardiac output monitoring device based on bioreactance technology which analyzes relative phase shifts of oscillating currents that occur when current traverses the thoracic cavity. Blood exiting the left ventricle to the aorta creates differences in the sensed current (time delay or phase shift), which NICOM measured to calculate the stroke volume. The sensors also contain an electrocardiogram element which measures heart rate and calculates CO which is reported as a 50 second average.^13^ The NICOM monitor is attached to a sphygmomanometer cuff which measured blood pressure at 10 minute intervals and allows for calculation of intermittent SVR.

### 3-dimensional transthoracic echocardiography (3DE)

Echocardiography-derived CO has been validated against the gold standard swan ganz thermodilution in pregnancy, and has been suggested as a reference for the validation of other CO techniques in pregnancy.^14^ Transthoracic echo was performed by qualified adult sonographers using Phillips iE33. Patients were scanned in the left lateral decubitus position. A three-lead electrocardiogram was used to enable image gating and record heart rate. Left ventricular (LV) ejection fraction (EF) was measured by 3D, which was reconstructed from apical three-, four-, and five-chamber views. CO was calculated directly by the Phillips machine on post-processing. Other measurements obtained included LV global strain and standard 2D and 3D measurements. (Table 3) 2D echocardiogram CO has inherent limitations due to angle-dependency of pulse wave Doppler and inaccuracies in left ventricular outflow tract size due to measurement inaccuracies and assumption of uniform morphology.^15^ 3DE eliminates these velocity and geometric assumptions, and also reconstructs CO over 7-14 cardiac cycles as opposed to a single beat which offers a broader view of flow quantification. Therefore, 3DE was chosen as the hemodynamic standard in this study. Kortakoff blood pressure (BP) obtained by a standard sphygmomanometer cuff was used as the reference standard for MAP.

### Data collection

Patients underwent 60 minute monitoring sessions with NICOM and CS at 2 timepoints: antepartum admission and postpartum day 1 or 2. Simultaneous 3DE was performed at the same time as NICOM and CS monitoring. Sphygmomanometer BP measurements were obtained at 10 min intervals.

### Statistics and Data Analysis

The primary outcome was CO (L/min). 3DE images for CO processing were acquired over a 10 minute period. Concomitant NICOM-CO and CS-CO during the same 10 minute interval of 3DE-CO image acquisition were averaged and used for comparison.

The secondary outcome was SVR (dynes*sec*cm^-5^). Using the same 10 minute time period as for the aforementioned CO measurements, concomitant average NICOM-SVR and CS-SVR were compared to 3DE-SVR. CS-SVR was measured directly by the device. For NICOM and 3DE SVR, the following equation was used: [(MAP – CVP)/CO] x80. For NICOM-SVR, NICOM-MAP and NICOM-CO were used in the equation. For 3DE, 3DE-CO and either NICOM-MAP or CS-MAP were used, and then compared to the same respective modality. CVP was derived from the echocardiographic estimate based on inferior vena cava diameter and respiratory variation.

Other hemodynamic parameters including SV (mL), HR (bpm), and (MAP) mmHg) were measured. For hemodynamic trend analysis, NICOM and CS data were averaged for each monitoring day. Echocardiographic measurements included LV EF, LV structure (wall thickness, systolic and diastolic volumes) and LV global longitudinal strain (GLS, %).

Data analysis was conducted utilizing IBM SPSS (version 20.0.0.0; 2021). Descriptive statistics were performed for the baseline demographics using frequency or median with interquartile range. With 3DE being considered the reference CO, we assessed for interchangeability with NICOM-CO and CS-CO. Interchangeability was assessed by correlation, accuracy (bias), and precision (95% limits of agreement [LOA]), and percentage error (PE).

Correlation was measured by simple linear regression and Pearson’s coefficient (R^2^), with 2 methods being considered interchangeable if R^2^ >=0.6. Bias and LOA were calculated with Bland-Altman analysis, and PE defined as 2 times the standard deviation of the bias over the mean.^16^ Two methods may be used interchangeably if the difference in bias is not clinically relevant and PE <=30%.^17^ For example, the average CO in normal pregnancy is 7.9 L/min by invasive monitoring^18^, so difference of up to 2L/min may be considered small and not clinically significant.

## Results

### Study population

Twelve patients were enrolled. Data from one patient was excluded due to poor echocardiographic acoustic windows. Therefore, eleven patients were analyzed and yielded 22 data sets.

From a maternal perspective, the median age was 33.5 years [29.5-35.3) and the median BMI at delivery was 35.5 kg/m^2^ [31.8-40.3]. At the time of enrollment, the median gestational age was 30 weeks [27.8-31.3] with median SBP 172 [167.3-181.8] and median DBP 98 [95.8-99.3]. Eight of the 12 patients required acute antihypertensive therapy for severe hypertension. Most commonly, patients received labetalol IV as the preferred therapy for acute severe BP management. From a fetal perspective, the median gestational age at delivery was 32 weeks [29.5-34]. Over half of the pregnancies were complicated by fetal growth restriction (66.7%) and nearly all neonates required neonatal intensive care unit admission (91.6%). There was one intrauterine fetal demise and no neonatal demises. (Table 1).

**Table 1:** Baseline characteristics of study population.

|  | **N=12 (%)** |
| --- | --- |
| **Maternal Age (y), median [IQR]** | 33.5 [29.5-35.3] |
| **Ethnicity** |  |
| Hispanic | 2 (16.6) |
| Non-hispanic | 10 (83.3) |
| **Race** |  |
| White | 8 (66.7) |
| Black | 1 (8.3) |
| Asian | 1 (8.3) |
| Other | 2 (16.6) |
| **Insurance** |  |
| Private | 9 (75) |
| Medicaid | 3 (25) |
| **Nulliparity** | 5 (41.6) |
| **BMI at delivery (kg/m<sup>2</sup>), median [IQR]** | 35.5 [31.8-40.3] |
| <25 | 1 (8.3) |
| 25-29.9 | 1 (8.3) |
| 30 to 34.9 | 3 (25) |
| 35-39.9 | 3 (25) |
| >40 | 4 (33.3) |
| **Pre-pregnancy BMI (kg/m<sup>2</sup>), median [IQR]** | 31 [27.8-34.5] |
| <25 | 1 (8.3) |
| 25-29.9 | 4 (33.3) |
| 30 to 34.9 | 4 (33.3) |
| 35-39.9 | 2 (16.6) |
| >40 | 1 (8.3) |
| **Gestational diabetes** | 1 (8.3) |
| **Smoking** | 2 (16.6) |
| **Chronic hypertension not requiring medications** | 1 (8.3) |
| **Gestational age at enrollment (weeks), median[IQR]** | 30 [27.8-31.3] |
| **Systolic blood pressure at enrollment (mmHg), median [IQR]** | 172 [167.3-181.8] |
| **Diastolic blood pressure at enrollment (mmHg), median [IQR]** | 98 [95.8-99.3] |
| **Antihypertensive management for severe hypertension** |  |
| Labetalol IV | 6 (50) |
| Nifedipine IR | 1 (8.3) |
| Hydralazine IV | 1 (8.3) |
| **HELLP syndrome** | 3 (25) |
| **Laboratory values upon hospital admission** |  |
| Urine protein: creatinine ratio, median [IQR] | 0.93 [0.3-4.6] |
| AST (U/L), median [IQR] | 32.5 [21.3-42.3] |
| ALT (U/L), median [IQR] | 21 [13-25.5] |
| Platelets (x1,000/microL), median [IQR] | 250 [174-272.8] |
| Creatinine (mg/dl), median [IQR] | 0.64 [0.6-0.7] |
| **Mode of delivery** |  |
| Vaginal delivery | 1 (8.3) |
| Cesarean delivery | 10 (83.3) |
| Dilation & evacuation | 1 (8.3) |
| **Gestational age at delivery (weeks), median [IQR]** | 32 [29.5-34] |
| **Fetal growth restriction** | 8 (66.7) |
| **NICU admission** | 11 (91.6) |
| **Neonatal death** | 0 |
| **Fetal death** | 1 (8.3) |
| **Birth-weight (g), median [IQR]** | 1300 [1110-1915] |
| **Apgar score at 1 minute of life, median [IQR]** | 6 [4-7.3] |
| **Apgar score at 5 minutes of life, median [IQR]** | 9 [8-9] |
| **Antihypertensive regimen upon hospital discharge** |  |
| None | 4 (33.3) |
| Labetalol | 1 (8.3) |
| Nifedipine | 4 (33.3) |
| Both nifedipine and labetalol | 3 (25) |

### Cardiac Output (Figure 1)

Pearson correlation coefficient comparing CS-CO with 3DE-CO showed strong correlation (R^2^=0.70, p=<0.001). Bland Altman analysis comparing CS-CO with 3DE-CO showed a mean bias +/- SD of 2.05 +/- 1.18 L/min, 95% LOA -0.20-4.31, and PE 29% error. Antepartum and postpartum CS calculations were similar. Postpartum CS-CO bias +2.1 +/-1.1 L/min with PE 28% and R^2^=0.72 (p = 0.002) versus antepartum CS-CO bias 1.9 +/- 1.2 L/min with PE 30% and R^2^=0.69 (p=0.002).

**Figure 1:**
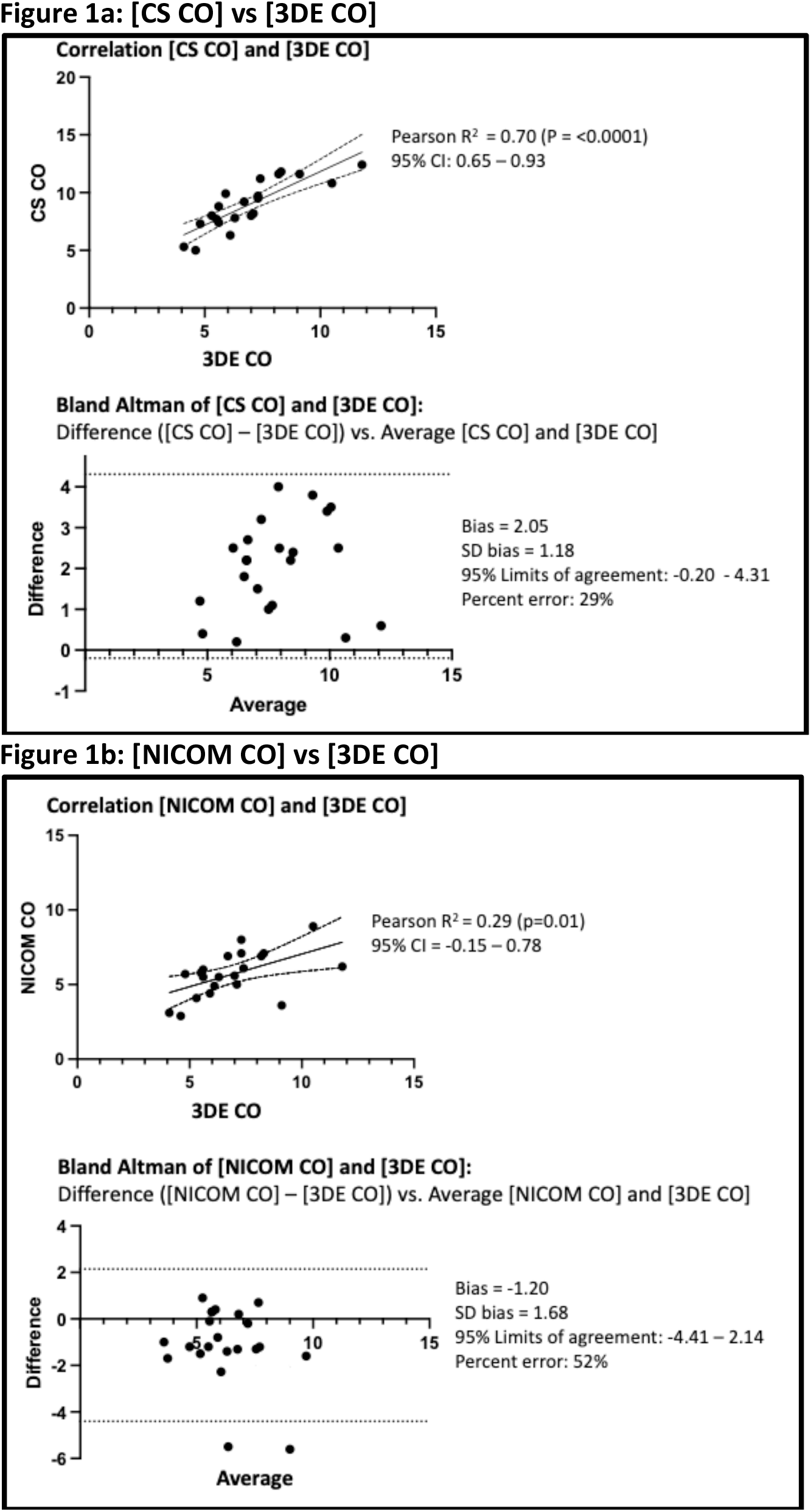
Cardiac Output: Correlation and Bland-Altman.

In contrast, Pearson correlation coefficient comparing NICOM-CO with 3DE-CO showed moderate correlation (R^2^=0.29, p=0.01). Bland Altman analysis comparing NICOM-CO with 3DE-CO showed a mean bias +/- SD of -1.2 +/- 1.68 L/min, 95% LOA -4.41 – 2.14, and PE 52%. High PE driven primarily by 2 outliers in the antepartum group. Comparing antepartum to postpartum NICOM measurements suggested better correlation and accuracy postpartum. Postpartum NICOM-CO bias -0.7 +/-1.74 L/min with PE 29% and R^2^=0.86 (p <0.001) versus antepartum NICOM-CO bias 1.18 +/- 4.18 L/min with PE 69% and R^2^= 0.27 (p=0.12).

### Systemic Vascular Resistance (Figure 2)

Pearson co-efficient comparing CS-SVR to 3DE-SVR showed strong correlation (R^2^=0.81, p=<0.0001). Bland Altman comparing CS-SVR to 3DE-SVR showed a bias +/- SD -279 +/- 156 dyes/sec/cm^5^, 95% LOA -580 – 18,4, and PE 28%.

**Figure 2:**
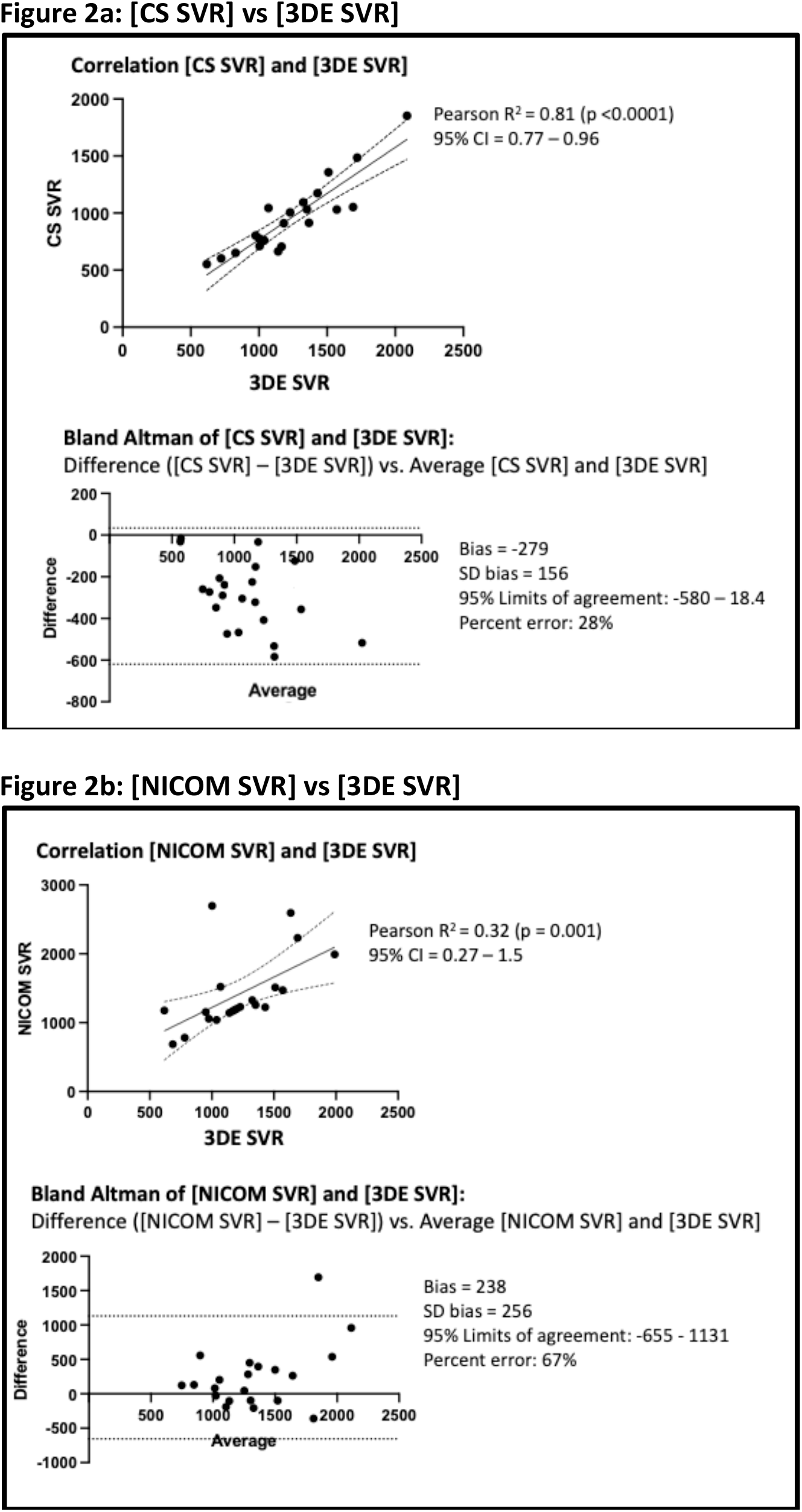
Systemic Vascular Resistance: Correlation and Bland-Altman.

Pearson co-efficient comparing NICOM-SVR to 3DE-SVR showed poorer correlation (R^2^=0.32, p=0.001). Bland-Altman comparing NICOM-SVR and 3DE-SVR CO showed a bias +/-SD 238 +/- 256, 95% LOA – 655 - 1131, and PE 67%.

### Mean Arterial Pressure

Pearson correlation coefficient comparing CS-MAP with sphygmomanometer BP showed strong correlation (R^2^=0.73, p=<0.01). Bland Altman analysis comparing CS-MAP with sphygmomanometer BP showed a mean bias +/- SD of 2.33 +/- 9.9 mmHg, 95% LOA -17 – 21.7, and PE 19%.

### Hemodynamic Trends (Table 2)

There was no significant difference between antepartum and postpartum CO, SVR, and MAP according to both modalities. However patient population was small, and these trends were not statistically significant.

**Table 2:**
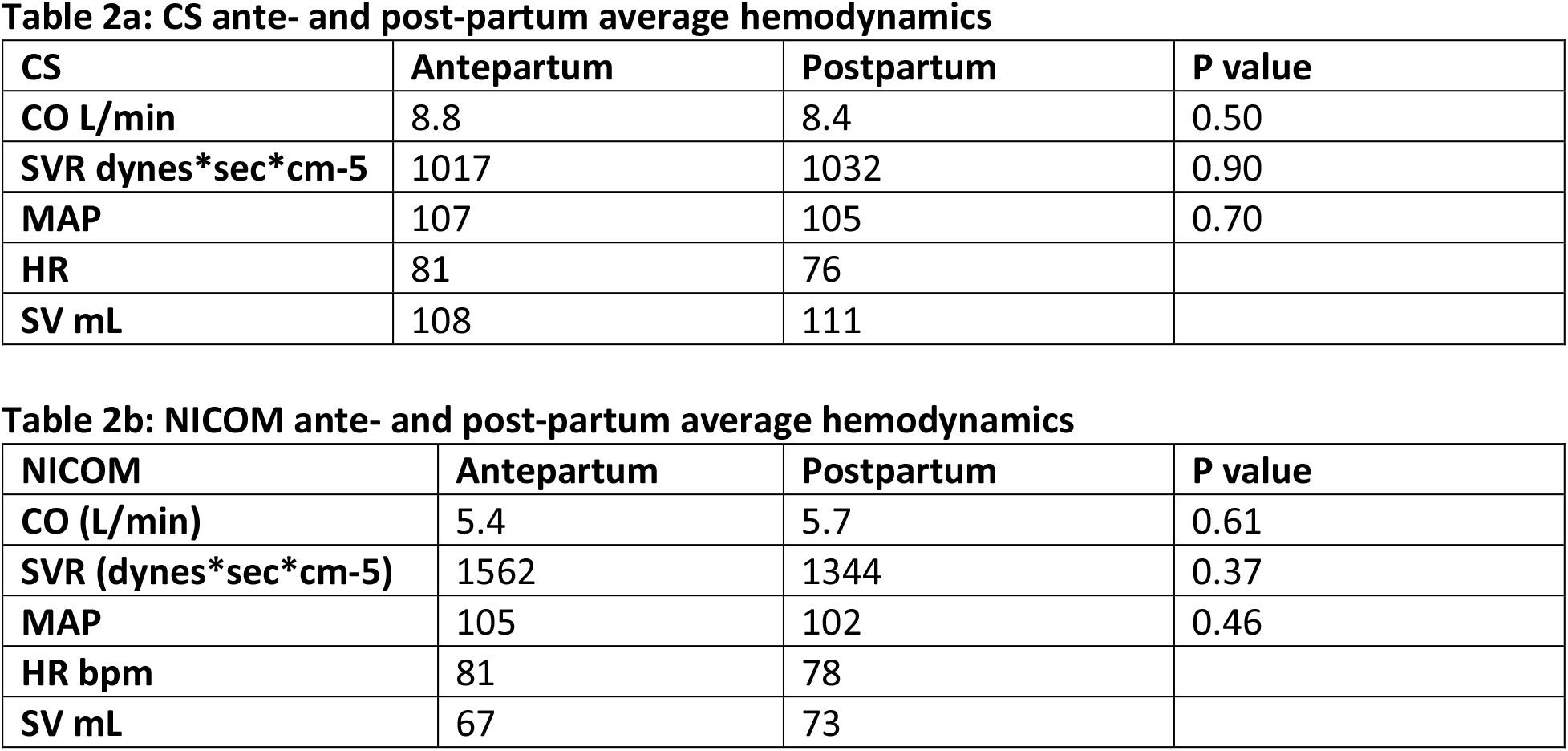
Average hemodynamics ante- and post-partum by modality: CS and NICOM.

**Table 3:** Echocardiographic Parameters ante- and post-partum.

|  | Antepartum | Postpartum |  |
| --- | --- | --- | --- |
| Parameter | Mean (SD) | Mean (SD) | P-value<br>(two sided) |
| CO (L/min) | 7.4 | 7.1 |  |
| SV (mL) | 92 | 88 |  |
| Left ventricular Ejection Fraction (%) | 63 (3) | 62 (5) | 0.68 |
| Global Longitudinal Strain (%) | -21.5 (1.5) | -20.2 (2.0) | 0.20 |
| Left ventricular End Diastolic Volume (ml) | 143 (41) | 159 (45) | 0.17 |
| Left ventricular End Systolic Volume (ml) | 53 (15) | 60 (19) | 0.14 |
| Interventricular Septal Diameter (cm) | 0.9 (0.2) | 0.9 (0.2) | 0.85 |
| Left ventricular posterior wall diameter (cm) | 1.0 (0.2) | 0.9 (0.2) | 0.53 |
| E/E' Average (cm/sec) | 8.2 (2.0) | 7.4 (2.0) | 0.58 |

### Echocardiographic Parameters (Table 3)

Between the antepartum and postpartum periods, there were no significant changes in left ventricular function or structure. The average EF was 63% antepartum and 62% postpartum. GLS was slightly lower postpartum (21.5% vs 20.2%), however this difference was not statistically significant, and both values were within normal limits.

## Discussion

We performed a prospective observational study in 12 patients with early onset severe preeclampsia to evaluate CO derived from the Clearsite Fingercuff and NICOM against 3DE-CO. To our knowledge, this is the first study evaluating both of these methods in the preeclampsia population. We found that CS-CO showed strong correlation and sufficiently low PE to be considered interchangeable with 3DE measurements. CS tended to over-estimate CO as compared to 3DE (bias +2.05 L/min). As previously described, two measurement methods may be considered interchangeably if the difference in bias is not clinically relevant and PE <=30%.^17^ Given that the average CO in normal pregnancy is 7.9 L/min by invasive monitoring^18^, our observed difference of approximately 2L/min may be considered small and not clinically significant. Furthermore CS-CO demonstrated strong correlation with 3DE-CO, with the difference between CS and 3DE rising as CO rises. This suggests that if needed for concordance, there is a systematic, linear offset for CS-CO that can be compensated for by applying a regression equation. CS-MAP correlated strongly and could be considered interchangeable with sphygmomanometer BP, which is consistent to prior studies.^19^ CS-SVR also had strong correlation and sufficiently low PE to be considered interchangeable with 3DE measurements. CS tended to under-estimate SVR as compared to 3DE (mean bias -293 dyes/sec/cm^5^), but may be considered small compared with the average pregnancy SVR of 980 dynes/sec/cm^5^.

Therefore while CS tends to slightly overestimate CO and underestimate SVR, these differences were small and not clinically significant. Therefore CS could be considered clinically interchangeable with 3DE for CO and SVR. These findings support Clearsite Fingercuff as a potential clinically useful modality for real-time hemodynamic monitoring in severe preeclampsia.

Our results differ from a prior study evaluating CS from 2DE TTE in 3^rd^ trimester pregnancy which showed an unacceptable agreement for CO measurements between these 2 modalities.^20^ This may be due to our use of 3DE as the reference standard, as opposed to 2DE. However given these contrasting findings, further studies to clarify the validity of CS in the preeclamptic population are warranted.

In contrast, NICOM- CO measurements showed only moderate correlation with 3DE-CO. Additionally, calculated PE was too high to meet criteria for interchangeability.^16^ NICOM-CO tended to under-estimate CO as compared to 3DE. While the lower bias suggests that NICOM- CO may be more accurate compared to CS-CO, the wider LOA suggests it is less precise.

Similarly, NICOM-SVR had poor correlation and a calculated PE too high to meet criteria for interchangeability. These findings suggest that NICOM would not be clinically useful for continuous hemodynamic monitoring in severe preeclampsia. Prior studies showed that NICOM CO assessment had acceptable correlation with echocardiogram-derived CO in normal pregnancy.^10,21^ Differences in our results could be explained by differences in body habitus – NICOM accuracy is limited by obesity, and the majority of our patients had BMI >30 kg/m^2^ at the time of delivery. This is in contrast to prior study cohorts with an average third trimester BMI of 26 kg/m^2^.^21^ The different hemodynamic profile in severe preeclampsia (lower CO and higher SVR as compared to normal pregnancy) may also be a factor, as Doherty et al found lower NICOM accuracy at lower CO.^10^

Interestingly, when comparing antepartum and postpartum NICOM-CO measurements against 3DE, NICOM demonstrated better accuracy and precision and lower PE in the postpartum period. It is also notable that the most outlying data points for NICOM-CO occurred during antepartum measurements. Thus NICOM inaccuracies in the antepartum period may be driven primarily by the gravid uterus elevating the diaphragm and altering thoracic anatomy. A prior study showed that upper abdominal surgical procedures reduced NICOM reliability compared to echocardiogram. Postulated reasons included upper diaphragm displacement which alter electrical flux through the thorax and changes in shape and position of the heart.

Similar physiologic changes occur during pregnancy, which would affect NICOM accuracy antepartum but resolve postpartum. Our findings suggest that while NICOM could be used for hemodynamic monitoring in the postpartum period, it cannot be considered clinically useful in the pregnant population at this time.

Our study showed no significant differences between antepartum and postpartum CO, SVR, and MAP for both the NICOM and CS modalities. These trends did not reach statistical significance due to the small study population; however, they are still hypothesis generating. After normal pregnancy, CO drops and SVR rises in the immediate postpartum period.^22^ The lack of change in CO and SVR in our severe preeclampsia population may be due to the effects of antihypertensive treatment. Since the majority of our patients received labetalol, this raises the possibility that CO may be higher in the postpartum preeclampsia population as compared to antepartum. Further studies are needed to clarify the hemodynamic effects of antihypertensives in severe preeclampsia, and if adjusting medical treatment based on hemodynamics can improve blood pressure control, hemodynamics, and maternal/fetal clinical outcomes.

There are several limitations to our study because it was performed at a single center with small sample size limited by disease selection and funding. First, meaningful concordance analysis was not feasible due to small sample size and limitation of one hemodynamic set per 3DE study. However correlation analysis suggests that CS-CO bias can be corrected with a regression equation, though this would not be feasible with NICOM-CO. Second, outlying data points, such as those seen in NICOM-CO, may have a more prominent effect on interchangeability analysis due to the small sample size. Larger studies in a similar patient population would help clarify the extent to which NICOM-CO accuracy is affected by anatomic limitations as described above, particularly in the antepartum period. Third, we utilized a reference method of 3DE for hemodynamics because invasive hemodynamics with a swan-ganz catheter was not feasible or safe in a pregnant population. Best efforts were made to ensure good acoustic windows and accurate hemodynamic values, but there still may be some discrepancy from invasive hemodynamic values. Lastly our study population represents a subset of the pregnant population, Therefore, applicability to other HDP populations, such as preeclampsia without severe features and gestational hypertension, requires further confirmation.

In summary, CS-CO and SVR could be considered be interchangeable with 3DE-CO and 3DE-SVR, respectively. CS tends to slightly overestimate CO and underestimate SVR, these differences were small and unlikely clinically relevant. Therefore, CS could be considered clinically interchangeable with 3DE for CO and SVR. In contrast, NICOM-CO and NICOM-SVR showed poorer correlation with less precision, and could not be considered interchangeable with 3DE-CO and 3DE-SVR, particularly in the antepartum period. These findings suggest that the CS fingercuff, but not NICOM, has potential to be clinically useful for real-time CO and SVR monitoring in patients with preeclampsia with severe features. Further studies in a larger sample size and in other hypertensive disorders of pregnancy are needed to confirm our findings and inform applications to clinical care.

## Data Availability

All data produced in the present study are available upon reasonable request to the authors.

## ABBREVIATIONS:^1^

^1^PEC: severe preeclampsia
CO: cardiac output
SVR: systemic vascular resistance
CS: Clearsite Fingercuff
NICOM: non-invasive cardiac monitoring with bioreactance
3DE3: 3D-echocardiography
PE: percentage error
LOA: limit of agreement
HDP: hypertensive disorders of pregnancy
SBP: systolic blood pressure
DBP: diastolic blood pressure
MAP: mean arterial pressures
SV: stroke volume
CVP: central venous pressure
LV: left ventricular
EF: ejection fraction
BP: blood pressure
GLS: global longitudinal strain

